# Multipoint Stimulation Motor Unit Number Estimation of the Extensor Indicis and Anconeus after Cervical Spinal Cord Injury

**DOI:** 10.1101/2024.08.08.24311675

**Authors:** Adenike A. Adewuyi, Mahdis Hashemi, Shreyaa Khanna, Michael J. Berger, Colin K. Franz

## Abstract

**Introduction:** The health of infralesional lower motor neurons (LMNs) after a cervical spinal cord injury is frequently overlooked, despite its critical role in mediating effective clinical interventions for improving arm and hand function. Prior studies suggest high frequencies of infralesional lower motor abnormalities in muscles that are potential targets for nerve transfer surgery, a procedure that has the potential to restore upper limb function.

**Methods:** In this prospective, two-center cohort study, we used multipoint stimulation motor unit number estimation (MPS-MUNE) to evaluate the number of motor units in clinically relevant infralesional muscles, including the predominantly C7-innervated anconeus and the predominantly C8-innervated extensor indicis (EI) in 15 individuals with cervical spinal cord injury (26 limbs) and 17 healthy controls.

**Results:** Both CMAP and MUNE values were significantly lower (p < 0.05) for those with cervical spinal cord injury (EI CMAP: 2.0 mV±1.57, EI MUNE: 33±30.5, Anconeus CMAP:2.7 mV±1.9, Anconeus MUNE: 39±50.6) versus controls (EI CMAP: 6.6mV±1.0, EI MUNE:137±33.9, Anconeus CMAP:6.6 mV±1.3, Anconeus MUNE: 146 ±42.3). The test-retest reliability as measured by intraclass correlation coefficient and confidence interval (CI) for the EI and anconeus were 0.84 (CI: 0.45-0.95) and 0.78 (CI: 0.36-0.93), respectively.

**Discussion:** This study shows significant loss of infralesional motor units after cervical spinal cord injury. We demonstrate the potential utility of MPS-MUNE for evaluating the health of LMNs. The LMN abnormalities observed underscore the significance of this approach to evaluating potential targets for nerve transfer surgery for the restoration of upper limb function.

## INTRODUCTION

Muscles innervated by the radial nerve and posterior interosseous nerve (PIN) are integral for elbow extension and grasp function, respectively. Recently, nerve transfer surgery has demonstrated promise in restoring function to these muscles in individuals with paralysis from cervical spinal cord injury (SCI).^1,2^ The restoration of elbow extension involves the coaptation of the axillary branch to the posterior deltoid or teres minor muscle above the neurological level of injury (typically supplied by the C5 myotome) to the radial nerve branches of the triceps in individuals with C5-C7 levels of injury. Similarly, hand opening function can be restored with the coaptation of the supinator branch (C5) to the infralesional PIN. Though the efficacy of nerve transfer is still a matter of debate, a recent study by Javeed *et al*. showed both statistical and clinically significant improvement in strength functional outcome measures after nerve transfer surgery in spinal cord injury ^2^. Given these novel surgical applications, robust electrophysiological measures of the radial and posterior interosseus innervated muscles are required.^3^ Recent studies have demonstrated abnormalities of the infralesional LMN that may impact an individual’s candidacy for nerve transfer surgery. However, these studies use non-quantitative methods such as the presence of denervation potentials (e.g. fibrillation potentials and positive sharp waves) which do not reflect the extent of LMN injury.^4,5^ For example, over time, denervation potentials may no longer be detected with milder LMN loss due to compensatory sprouting and muscle reinnervation by surviving motor neurons. Alternatively, in severe LMN injury, chronic denervation can lead to muscle fibrosis and the reduction of spontaneous electrical activity. This is a clinically meaningful distinction since fibrosed muscles that no longer demonstrate these denervation potentials are not good candidates for nerve transfer procedures.

Motor unit number estimation (MUNE) has long been used by clinicians and researchers to develop a more detailed and quantitative understanding of the neuromuscular system in a host of physiological and pathological conditions, with the basic premise that the compound muscle action potential (CMAP) alone does not provide enough information about motor unit loss or remodelling over time.^6-8^ Although there are many MUNE techniques available, the most accessible to clinicians is the original incremental technique and its subsequent derivative techniques, including the original and adapted multiple point stimulation methods (MPS). These techniques require little additional expertise or specialized equipment but are limited by several technical pitfalls.^9^ MPS requires access to superficial nerves with long courses providing multiple stimulation sites. Furthermore, only a select few distal muscles are amenable to study with MPS, as proximal muscles have less “real estate” for surface nerve stimulation and more proximal muscles are prone to volume conduction error, whereby coactivation of muscles with the same nerve supply leads to overestimation of MUNE.^10^ Therefore, MUNE with MPS has typically been obtained only for the thenar and hypothenar muscle groups in the hand, as well as the extensor digitorum brevis in the foot. ^11-13^

The radial motor study of the anconeus is rarely performed in routine clinical settings and MUNE studies of this muscle are limited. Despite its small size, it is a useful muscle for electrodiagnostic investigation of neuromuscular conditions.^14-16^ Moreover, it often receives the same innervation as the lateral head of triceps and as it is small and more distal, the anconeus is more amenable to evaluation with the MPS-MUNE technique. Conversely, the radial motor study to extensor indicis (EI) is commonly performed in clinical settings and provides a reliable CMAP. However, to our knowledge, MPS-MUNE for this muscle has not been previously evaluated. Our primary objective was to evaluate and establish differences in MPS-MUNE estimates between individuals with SCI and controls. Further, we describe an approach for MPS-MUNE in the EI and establish test-retest reliability for both the EI and anconeus.

## METHODS

Neurologically intact control subjects and individuals with SCI were recruited at the Shirley Ryan Abilitylab in Chicago, IL, USA and the International Collaboration on Repair Discoveries (a.k.a. ICORD) at the University of British Columbia, Vancouver, British Columbia, Canada. All subjects gave their informed written consent, and the study protocol was approved by Institutional Review Boards at both institutions. Inclusion criteria for the SCI group included (1) people ages 18-80 with subacute (< 1 year and > 3 weeks from injury) or chronic (≥1 year from injury) injuries and (2) cervical neurological levels of injury at C1-C8 and (3) no concurrent peripheral nerve conditions unrelated to their spinal cord injury (i.e., brachial plexopathy or peripheral neuropathy). Inclusion criteria for the control group included people ages 18-80 without any neurological conditions. Routine clinical sensory nerve conduction studies of the radial, median and ulnar nerves were performed to rule out any superimposed peripheral neuropathy in the SCI group.

### Experimental Setup

The anconeus and EI of the dominant side for each control subject were evaluated. For the SCI group, bilateral anconeus and EI were evaluated, unless superimposed peripheral neuropathy or brachial plexopathy precluded evaluation. Skin temperature was maintained above 32^°^ C during the nerve conduction studies. The subjects were supine on an examination bed with shoulder adducted, forearm in prone position, and elbow flexed 90^°^ for anconeus evaluation. The active recording electrode was placed over the motor point of the anconeus (approximately 2 cm distal to the olecranon and 1 cm lateral to the border of the ulna) and the reference electrode was placed on the ulnar styloid. The ground electrode was placed between the recording electrode and stimulator, just proximal to the antecubital fossa. All data were collected using either Cadwell Sierra Summit (Shirley Ryan AbilityLab) or Natus Ultrapro (UBC/ICORD) EMG Systems. The commonly used clinical nerve conduction montage was used for the EI. The active recording electrode was placed over the motor point of the EI (about 4 cm proximal to ulnar styloid and 1cm radial to the border of the ulna), the reference electrode was placed on the ulnar styloid and the ground electrode was placed on the volar forearm in between the recording electrode and the stimulating electrode.

### MPS-MUNE

A supramaximal percutaneous stimulation was delivered to the radial nerve or PIN, 8cm proximal to the recording electrode by a handheld bipolar stimulator to record the supramaximal CMAP of the anconeus and EI, respectively. Ten surface motor unit potentials (SMUPs) with different amplitudes, sizes and shapes were recorded by gradually adjusting the stimulus intensity to produce an all-or-none response. Each SMUP was generated by moving the stimulator along the radial nerve and slightly altering the orientation of the stimulator until a SMUP was elicited above its threshold. There was no minimum amplitude prespecified as the lowest acceptable one. Motor potentials with identical morphology, amplitude, and onset there were reliably elicited at least 3 times were identified as SMUPs. In cases of severe atrophy, with less than 10 SMUPs, as many SMUPs as possible were recorded. The MUNE value was calculated by dividing the supramaximal CMAP amplitude by the mean amplitude of the 10 SMUPs. Fifteen control subjects attended additional testing sessions to evaluate the test-retest reliability of the MPS for both the anconeus and EI. These recording sessions were separated by a minimum of 24 hours. Figure 2 depicts the experimental setup with sample recordings of the CMAP and SMUP for a spinal cord injury subject.

**Figure 1:**
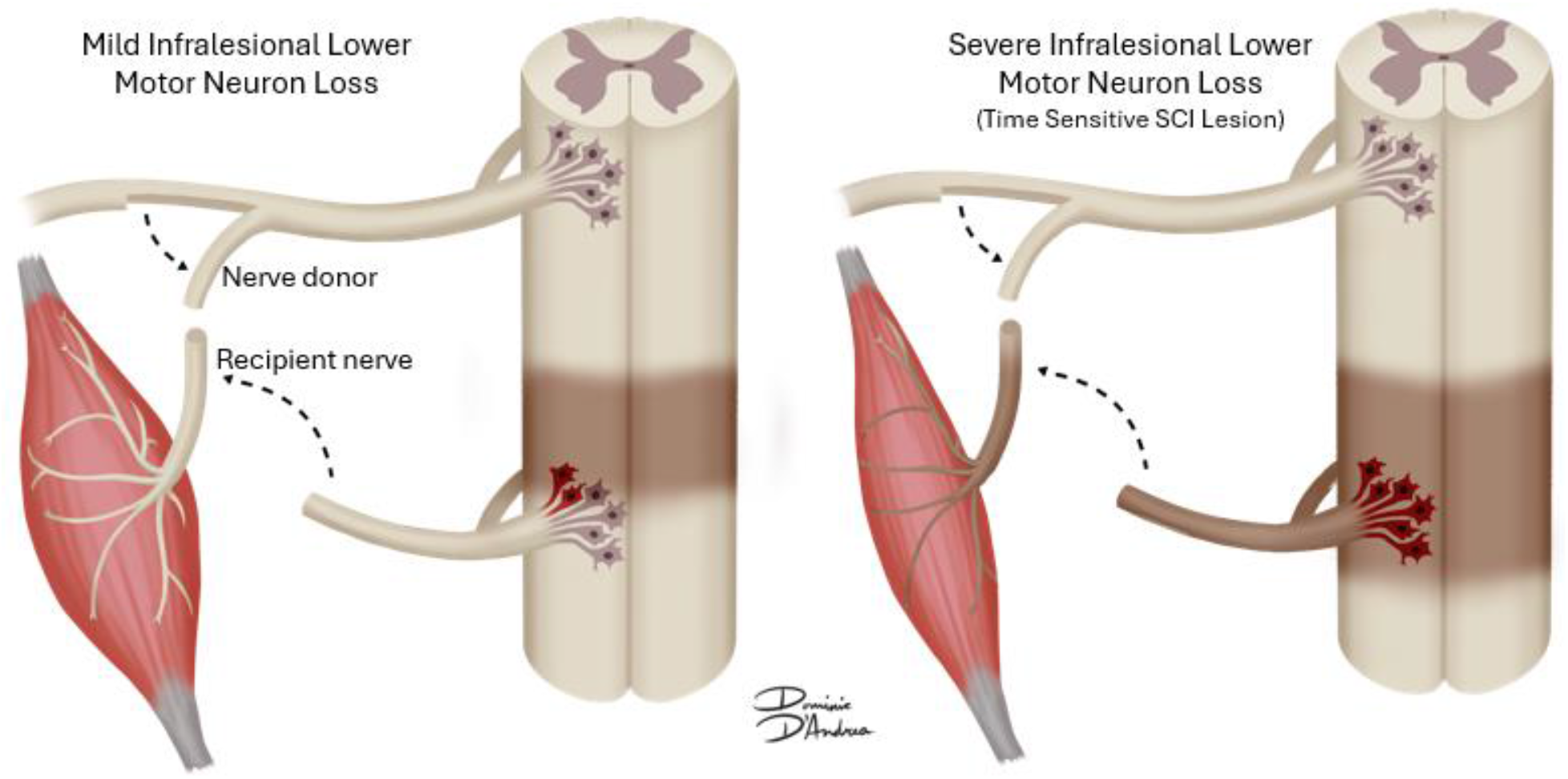
Role of nerve transfer in spinal cord injury. In nerve transfer surgery, restoration of function is achieved by coapting a supralesional donor nerve to an infralesional recipient. Healthy recipient infralesional motor neurons promote more favorable surgical outcomes. Whereas more extensive infralesional lower motor neuron loss may necessitate earlier intervention before irreversible degeneration atrophy occurs.

**Figure 2:**
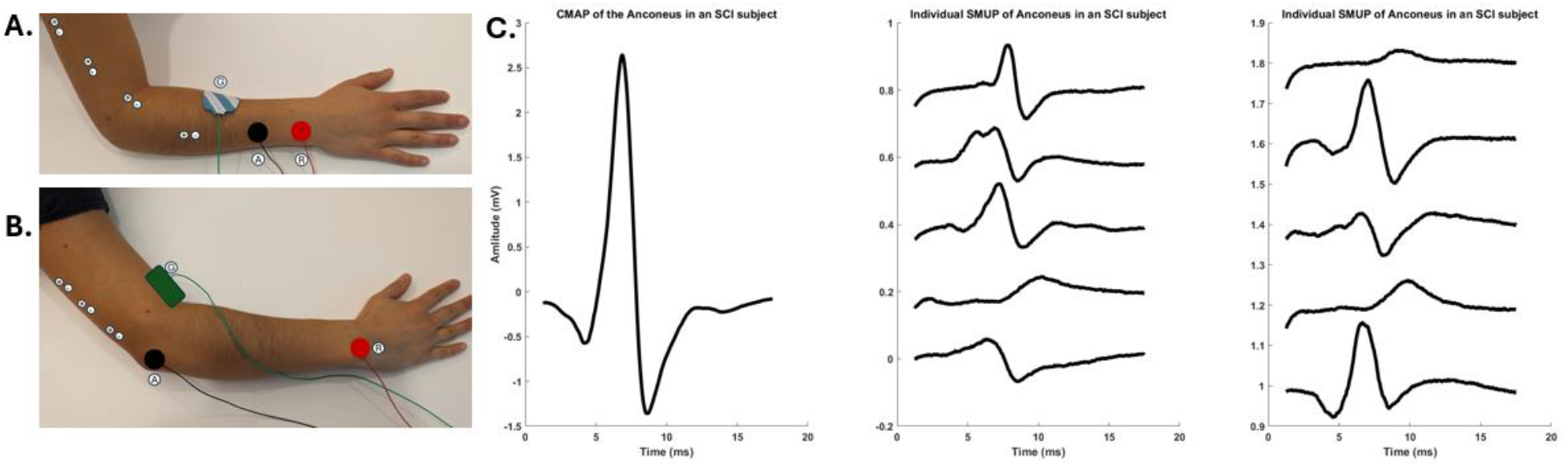
Experimental Setup: A. Electrode configuration for extensor indicis. B. Electrodes configuration for the anconeus. C. Sample compound muscles action potential (Left panel) and single motor unit action potentials (right two panels) elicited from the anconeus of an individual with a spinal cord injury.

### Statistics

The data from the test–retest protocol were examined graphically with Bland–Altman plots^17^ to determine if error was evident. The Shapiro-Wilk test was used to test assumption of normality. Test–retest reliability was examined with intraclass correlation coefficient ICC (2,1) and 95% CI, using Matlab (Mathworks R2023b). A one-way analysis of variance (ANOVA) was performed with experimental group as a fixed effect and significance level was set at 0.05.

## RESULTS

Fifteen individuals with SCI (14 males, 1 female, median age = 28, range = 18-65) and seventeen control subjects (10 males, 7 females, median age = 21, range = 20-43) with no known neurological deficits, were evaluated (Table 1). Fourteen subjects in the SCI cohort had traumatic injuries and eleven subjects had motor complete injuries (Table 2). Ten subjects had subacute SCI and five subjects had chronic SCI.

**TABLE 1:** Demographic Characteristics for Subjects.

| Basic Characteristics | Control<br>n = 17 | SCI<br>n = 15 |
| --- | --- | --- |
| **Sex:** |  |  |
| Male | 10 | 14 |
| Female | 7 | 1 |
| **Age:** Median (Range) | 21 (20-43) | 28(18-65) |
| **Time Since Injury** |  |  |
| Subacute | --- | 10 |
| Chronic | --- | 5 |
| **Injury Mechanism:** |  |  |
| Traumatic | --- | 14 |
| Nontraumatic | --- | 1 |

SCI: Spinal Cord Injury Cohort

**TABLE 2:** Neurologic Injury Classification for the Spinal Cord Injury Cohort.

| SCI Classification (# of subjects) |  |
| --- | --- |
| **NLI** |  |
| C1-C4 | 7 |
| C5 | 4 |
| C6 | 3 |
| C7 | 0 |
| C8 | 1 |
| **Motor Complete** | 11 |
| **Motor Incomplete** | 4 |

NLI: Neurologic Level of Injury, SCI: Spinal Cord Injury

### Test-Retest Reliability

Figure 3 (left panel) shows a regression line between the estimated number of motor units for two different trials for control subjects. The right panel shows Bland Altman graphs where the difference between the two trials is plotted against the mean estimated number of motors units from both trials. There was no observed systematic bias between trials. The differences between trials were normally distributed (*p* = 0.99 for the anconeus and *p* = 0.31 for the EI). The test-retest reliability for the anconeus was ICC=0.78 (CI: 0.36-0.93) and for the EI was ICC=0.84 (CI: 0.45-0.95).

**Figure 3:**
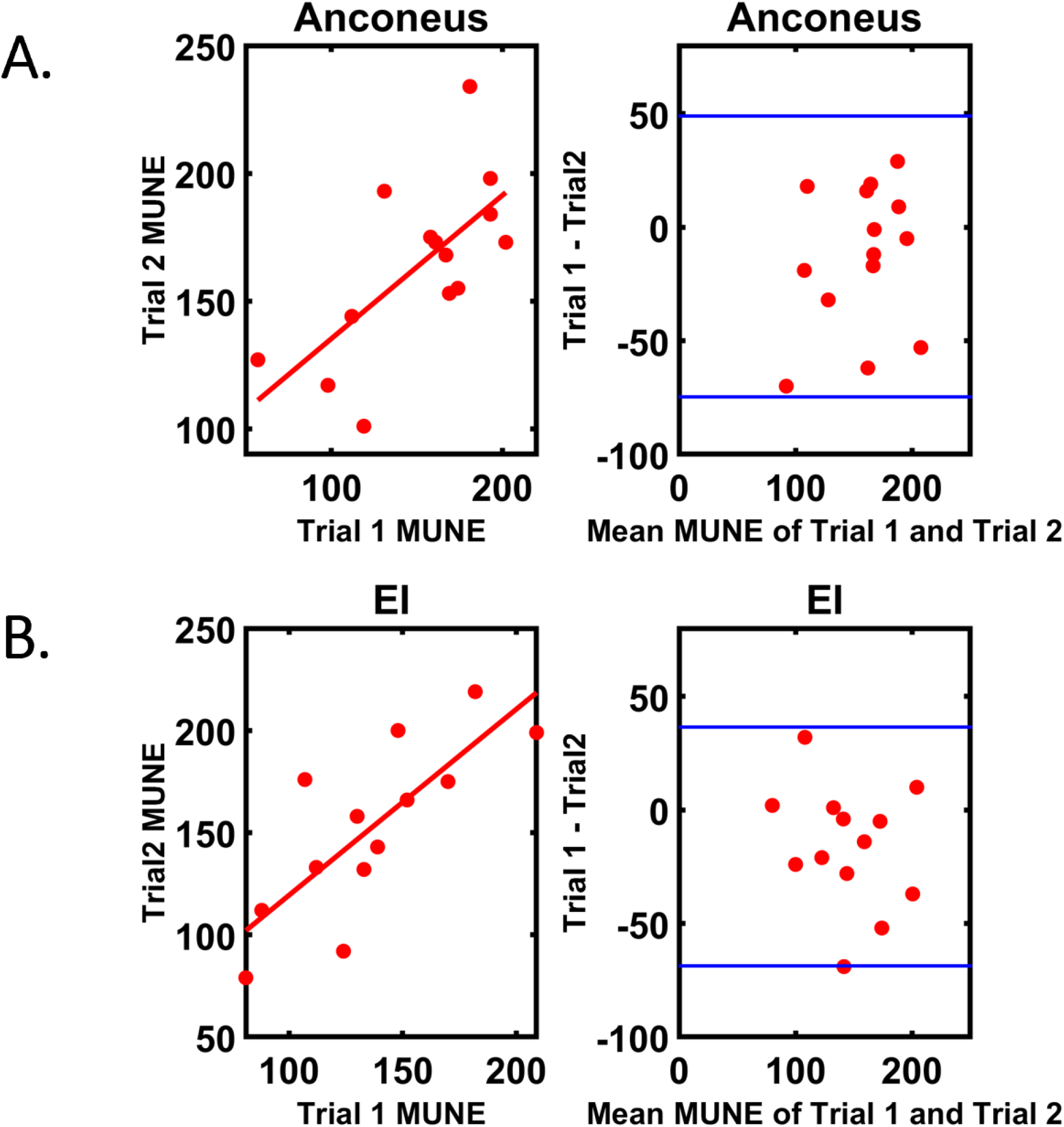
**Left**: Regression line between two different trials of MUNE calculations performed at least 24 hours part in control subjects. Bland-Altman Plots (right) of the difference between two different trials of MUNE calculations. Blue lines present 95% confidence interval. Fig3A: Anconeus. Fig3B: Extensor Indices (EI).

The mean anconeus CMAP for the control and SCI groups were 6.6 mV (± 1.28) and 2.7 mV (± 1.88) respectively. The mean anconeus MUNE for the control and SCI groups were 146 (±42.3) and 39 (± 50.6) respectively (Figure 4). Of note, one individual in the SCI cohort who represents the outlier for the SCI anconeus MUNE data in Figure 4B had marked clinical asymmetric weakness and initially presented with a Brown-Sequard-like syndrome after a motor vehicle collision. Only his left (stronger) side was included in this study as there was concern for superimposed brachial plexopathy on the right. The mean EI CMAP for the control and SCI groups were 6.6 mV (± 0.98) and 2.0 mV (± 1.57) respectively. The mean EI MUNE for the control and SCI groups were 137 (±33.9) and 32.7 (± 30.5) respectively (Figure 4). The differences in CMAP and MUNE estimates for both the anconeus and EI between controls and the SCI cohort were statistically significant (p<0.05).

**Figure 4:**
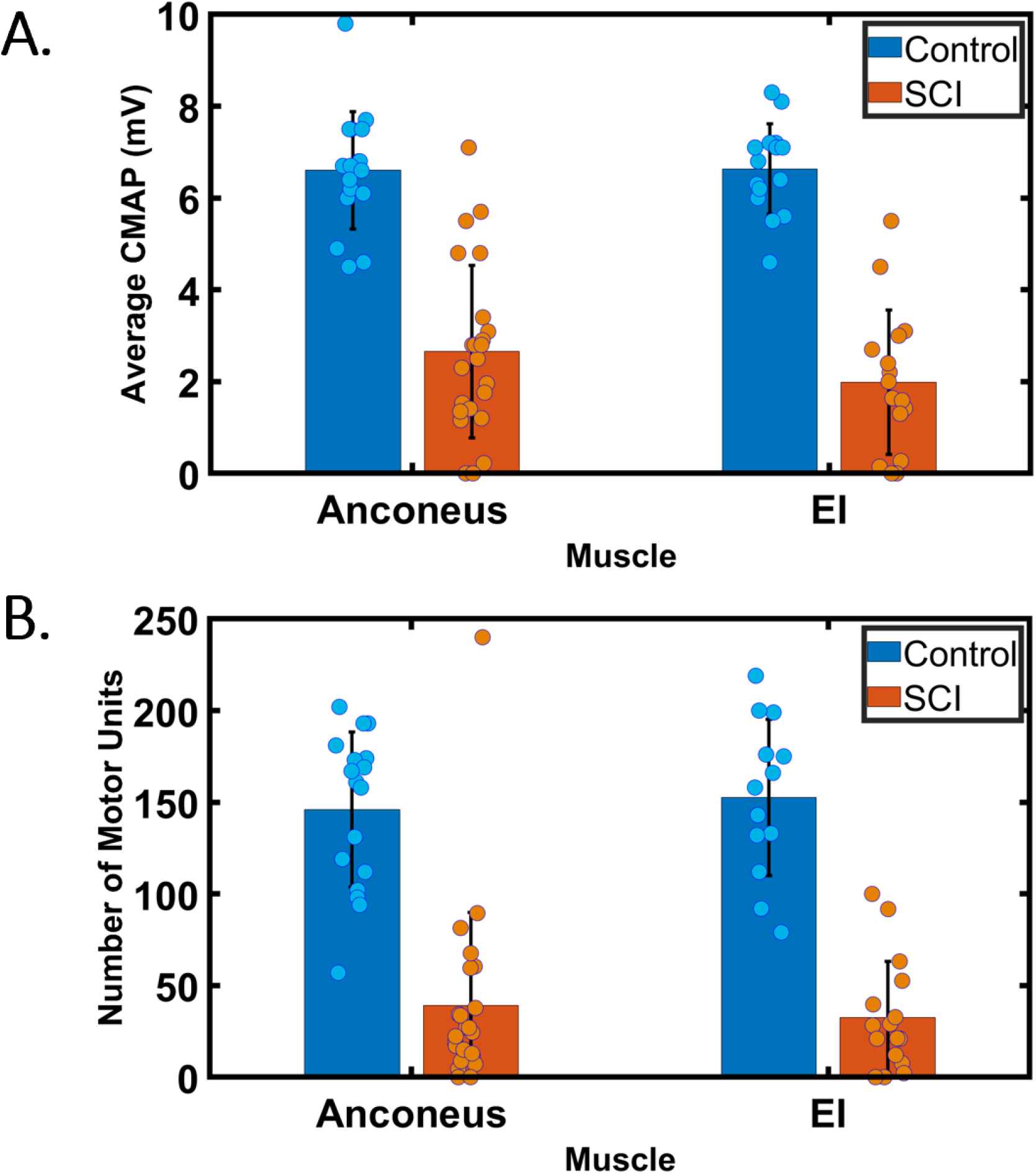
Comparison of the (A) compound muscle potential (CMAP), and (B) estimated number of motor units between controls and individuals with cervical spinal cord injury (SCI) for the anconeus and Extensor Indicis (EI). There is a significant difference (*p* < 0.01) between controls and SCI groups for both the anconeus and EI.

## DISCUSSION

This study demonstrated that the CMAP amplitudes and MUNE estimates of the anconeus and EI in individuals with SCI is significantly lower than controls. The results also demonstrated good test-retest reliability for the anconeus and EI. As the PIN and radial nerve are essential for wrist extension and hand opening, they are common recipients in nerve transfer procedures. Consequently, the health of these nerves has implications for nerve transfer candidacy. To our knowledge, there have been no prior MUNE EI studies in either controls or SCI. In a previous analysis of a cohort with chronic cervical SCI, we showed evidence of lower motor neuron abnormalities in 87% of individuals in muscles that are typical recipients for nerve transfer surgery.^5^ We have also shown in a recent retrospective study, a high prevalence of lower motor neuron abnormalities early after cervical SCI, below the neurological level of injury, using needle electromyography to document abnormal spontaneous activity. However, as abnormal spontaneous activity is non-specific and not quantitative, its pathophysiological significance is unclear. Our data not only shows abnormally low CMAPs but loss of motor units to infralesional C7 and C8 muscles that is not completely explained by the low CMAPs. These findings were observed in individuals with subacute injures as early as 1-3 months after injury which has implications for the timing of a nerve transfer procedure, since most recipient muscle targets for hand opening and hand closing receive nerve supply from C7-C8 spinal segments.

The mechanism of loss of infralesional motor units after SCI has yet to be elucidated. Though direct injury to the lower motor neuron would certainly cause loss of motor units, it may not explain loss of motor units in C7 and C8 innervated muscles in individuals with injuries at the C6 level and higher. Trans-synaptic degeneration, describes the hypothesis that a loss of supralesional input contributes to infralesional degeneration of LMN and muscle changes.^18^ However, the time course of this potential degenerative process is unclear and it is uncertain whether trans-synaptic degeneration could explain the loss of units within 3 months after an injury. Van De Meent et al evaluated the CMAP of the abductor digiti minimi and the abductor hallucis over time within the first year after a cervical SCI and found reduced CMAP amplitudes 2-4 weeks after injury with improvement in CMAP 6-8 months after injury.^19^ The initial reduction of CMAP amplitude is suggestive of motor unit loss followed by collateral reinnervation. Xiong et al similarly found low CMAP amplitudes and MUNE of the tibialis anterior in those with complete motor, subacute, cervical to thoracic SCI which significantly improved after 1-3 months.^13^ An important consideration is that the direct trauma to the cord may extend several segments beyond the clinical neurological level of injury in a longitudinal injury and as such may not correlate with clinical neurological level of injury. Further, no studies have correlated these EMG findings to spinal MRI imaging which would provide additional insight. It would also be of use to study the natural course of lower motor neuron loss after a SCI with serial MUNE studies as this would also be beneficial for planning interventions such as nerve transfer surgeries.^20^

Our anconeus MUNE estimates for control subjects are higher than what has been previously reported in the literature. Stevens *et al*. determined an anconeus MUNE estimate of 25-58 in healthy subjects using decomposition enhanced spike-triggered averaging (DE-STA), which is considerably lower than our estimate of 146. This is presumably related to the methodology of DE-STA which requires voluntary activation at submaximal contraction levels of 20-50%. Gilmore *et al*. had similar findings with DE-STA when comparing anconeus MUNE estimates of young and older subjects. As DE-STA employs high contraction levels of up to 50%, this may lead to oversampling of large units and a high chance of superimposition of multiple units leading to an underestimation of the MUNE. More recently, Zong et al, used MScanFit for MUNE estimation of healthy control subjects which also demonstrated low MUNE of 55. MScanFit is a recently developed MUNE method based on the CMAP scan and a model simulation of the responses. In MScanFit, a size limit of 25uV is set to prevent the addition of inappropriate units by the fitting procedure. As no size limit was set in this study and units as small as 10uV were observed in healthy controls, this may explain the discrepancy in MUNE estimates. Moreover, MScanFit appears to have lower estimates of MUNE in all muscles when compared to other techniques.^11,21,22^ Ultimately, further studies evaluating different MUNE methods in these previously understudied muscles are needed.

### Limitations

This study had a heterogeneous group of individuals with SCI and included multiple neurological levels of injury and variable time since injury which may explain the variability in the SCI results. Much of the SCI cohort were young men in their 30s or younger and as such, we cannot comment on the potential role of age and/or sex as modifiers of LMN injury. Finally, as LMN abnormalities were observed in both subacute and chronic SCI, a larger study with serial MUNE in those with acute/subacute and chronic SCI would yield significant insights regarding the temporal process of LMN injury recovery after SCI.

## CONCLUSION

The findings of this study shed light on the potential utility of MPS-MUNE for evaluating the health of lower motor neurons in individuals with cervical SCI, which has not been widely explored before. This study introduced the application of MPS MUNE to evaluate the posterior interosseous innervated EI muscle, expanding the range of muscles that can be assessed using this technique. We demonstrate the reliability of MUNE in clinically important muscles and show dramatic loss of infralesional motor units after SCI. The observed motor unit abnormalities in muscles that are potential targets for nerve transfer surgery indicate the significance of this approach for informing clinical interventions aimed at restoring upper limb function. It is noteworthy that the exact mechanisms underlying the observed motor unit abnormalities in SCI individuals remain to be fully elucidated.

While direct lower motor neuron injury is a potential factor, the concept of trans-synaptic degeneration and other factors might contribute to these alterations. Future studies that quantitatively track MUNE changes with serial assessments after SCI will be valuable for understanding the temporal dynamics of these changes and for guiding the timing of interventions like nerve transfer surgeries.

## Data Availability

All data produced in the present study are available upon reasonable request to the authors.

## Acknowledgement

A.A.A. would like to acknowledge support from the Foundation for Physical Medicine and Rehabilitation (Richard S. Materson ERF New Investigator Research Grant). C.K.F. would like to acknowledge support from the Belle Carnell Regenerative Neurorehabilitation fund. M.J.B. would like to acknowledge support from Wings for Life Project Grant (GR024775).

